# Longitudinal personal environmental exposure and *Streptococcus pneumoniae* colonisation in a cohort of children 5-14 years old in rural Gambia

**DOI:** 10.64898/2026.10.08.26364987

**Authors:** Dam Khan, Brenda Kwambana-Adams, Peggy-Estelle Tientcheu, Shola-Able Thomas, Peter Sylvanus Ndow, Ma-Ansu Kinteh, Archibald Worwui, Nuredin Ibrahim Mohammed, Mark P. Nicol, Martin Antonio

## Abstract

**Background:** Air pollution contributes substantially to morbidity and mortality in low-income countries, where reliance on solid fuels leads to high levels of household exposure. Colonisation of the upper respiratory tract by *Streptococcus pneumoniae* is a prerequisite for pneumococcal disease, and inhaled air pollutants may influence colonisation through impaired mucociliary clearance. Therefore, we investigated the association between *S. pneumoniae* colonisation and personal exposure to particulate matter (PM_2.5_) and carbon monoxide (CO) as well as temperature and relative humidity in older children in The Gambia.

**Methods:** We conducted a prospective study of 134 children aged 5-14 years across eight visits in rural Gambia. Personal 24-hour PM_2.5_, CO, temperature and relative humidity exposures were measured using personal monitors. Concurrently, combined nasopharyngeal/oropharyngeal swabs were collected at each visit, and *S. pneumoniae* colonisation was determined by microbiological culture. Mixed-effects logistic regression models were used to examine associations between environmental exposures and colonisation. Exposures were additionally decomposed into between-child and within-child components. Models were adjusted for age, season, and household income tertile.

**Results:** A total of 554 child visits with paired exposure and microbiological data were analysed. Overall, 24-hour geometric mean PM_2.5_ and CO were 12.6 µg/m³ (95% CI 12.2–13.1) and 0.16 ppm (95% CI 0.14–0.19) respectively and overall *S. pneumoniae* colonisation prevalence was 39.7%. In conventional models, PM_2.5_, CO, temperature, and relative humidity were not significantly associated with *S. pneumoniae* colonisation after adjustment for confounders. In the within-between decomposition analysis, higher between-child temperature exposure was consistently associated with lower odds of colonisation (mutually adjusted model: OR 0.74, 95% CI 0.56–0.98; p = 0.034). Relative humidity showed an inverse within-child association in crude analysis (OR 0.70, 95% CI 0.52–0.93; p = 0.013), which attenuated after adjustment for season and other confounders. No associations were observed for PM_2.5_ or CO exposures.

**Conclusion:** At the exposure levels observed in this cohort of older children, PM_2.5_ and CO exposures were not independently associated with *S. pneumoniae* colonisation. The pattern observed for temperature suggests that longer-term temperature conditions may be more relevant to pneumococcal colonisation than short-term exposure variation in this setting.

## Background

Air pollution is a major extrinsic driver of global morbidity and mortality rates. In 2019, it was responsible for an estimated 1.1 million deaths across Africa, with more than half of the fatalities attributable to household air pollution (1). In sub-Saharan Africa, household air pollution is primarily derived from the open combustion of biomass fuel or the use of inefficient stoves for cooking and heating. This process generates high emissions of harmful pollutants, including particulate matter with aerodynamic diameters ≤2.5µm (PM_2.5_) and carbon monoxide (CO). Although access to clean household energy has improved globally over the past two decades, from 51% in 2003 to 74% in 2023, Africa has seen minimal gains, with over 75% of households in sub-Saharan Africa still relying on polluting fuels (2). Acute respiratory infections (ARIs) are among the major health outcomes linked to air pollution, with increased PM_2.5_ exposure associated with higher rates of emergency department visits, hospital admissions and mortality from pneumonia (3–5). *Streptococcus pneumoniae* is a leading bacterial cause of pneumonia morbidity (6), and upper respiratory tract colonisation is a prerequisite for both pneumococcal disease and transmission (7, 8). Exposure to air pollutants has been shown to impair mucociliary clearance by slowing ciliary beating, disrupt airway epithelial barrier integrity, and alter innate responses (9–11), all of which may facilitate *S. pneumoniae* colonisation of the respiratory tract (12, 13). Despite these plausible biological mechanisms, epidemiological evidence linking air pollution to *S. pneumoniae* colonisation remains limited, particularly in older children (5-14 years old). This age group is epidemiologically important because of frequent social mixing and its role in transmitting *S. pneumoniae* to younger and more vulnerable children who bear the greatest burden of pneumococcal disease (14). Understanding how air pollution influences colonisation in older children may inform strategies to reduce *S. pneumoniae* transmission and LRTIs at the population level.

Most studies examining the relationship between air pollution and *S. pneumoniae* colonisation have relied on indirect air pollution exposure estimates, which may not accurately reflect individual exposure. Although recent studies have incorporated personal exposure measurements, these have largely focused on infants with few repeated measurements or been incorporated in intervention-based settings, thereby limiting generalisability (15–17). As a result, they do not capture within-child variability in exposure and colonisation over time, particularly beyond early life. Given that air pollution exposure varies across environments, behaviors, and socioeconomic contexts, more studies are needed that integrate repeated personal exposure measurements with longitudinal assessments of colonisation in older children.

In this study, we leveraged repeated measurements of personal exposure to PM_2.5_, and CO, as well as temperature and relative humidity in a cohort of healthy older children in The Gambia to address this knowledge gap. We examined whether personal exposure to PM_2.5,_ CO, temperature, and relative humidity was associated with *S. pneumoniae* colonisation. We also reported variations in levels of PM_2.5,_ and CO by household and environmental factors.

## Methods

### Study design and participants

This longitudinal, observational, prospective study was conducted in rural communities in the Foni Bintang-Karanai District in the western region of The Gambia (Figure 1). The villages in this district primarily rely on subsistence farming for their livelihoods and generally live in extended family compounds sharing a central courtyard where activities, including cooking and socialising take place. Villages typically have a population of between 200 and 1000 people.

**Figure 1:**
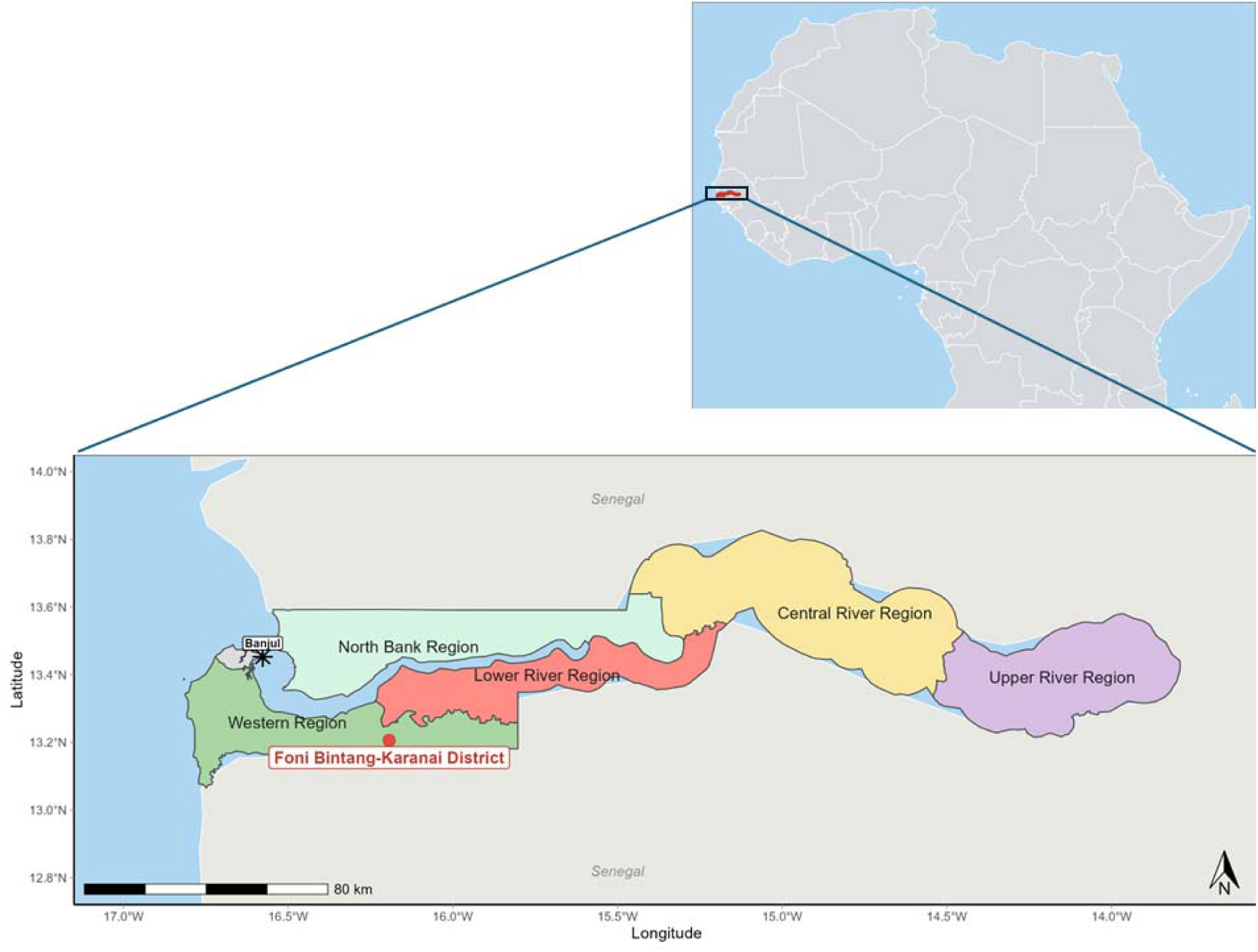
Map of The Gambia. The red dot represents the study area.

Following community engagement and sensitisation programmes, 134 healthy children aged 5 to 14 years were enrolled in the study. The exclusion criteria included participants with pre-existing medical conditions for which they were receiving antibiotic treatment and those who permanently relocated outside the study area during follow-up. The enrolled children were followed across eight study visits conducted at three-month intervals. Field activities for the first four visits were conducted from February 2019 to March 2020. Field activities were suspended between April 2020 and January 2021 due to COVID-19 mitigation measures. The next four visits were conducted between February 2021 and March 2022. Data on health and demographic covariates were recorded via face-to-face surveys at each visit using the REDCap software on electronic tablets. Study participants were asked questions on household and environmental factors including primary cooking fuel type, presence of a smoker in the household compound, and distance from the compound to the nearest main road.

### PM_2.5_, CO, temperature and relative humidity exposure measurements

Personal exposure to PM**_2.5_**, CO, temperature, and relative humidity was measured using the Particle and Temperature Sensor (PATS+) personal monitor (18). Minute-level PM_2.5_ concentrations (nephelometer-based, expressed in μg/m³), CO concentrations (electrochemical sensor, expressed in ppm), ambient temperature (°C), and relative humidity (%) were recorded. The lower detection limit (LDL) of PATS+ for PM_2.5_ is 10 μg/m³, and the upper detection limit is 30,000–50,000 μg/m³. The measurement range for CO is 0-500 parts per million (ppm). The device has a temperature measurement range of 0–50°C and relative humidity range of 0–95%. The devices were configured and calibrated using the PATS+ Platform for Integrated Cookstove Assessment (PICA) system software and following manufacturer’s protocol.

Field workers deployed the devices in the participants’ homes at each visit. The devices were placed in a waterproof custom-designed holder, and each participant was asked to wear the device strapped to their waist for a continuous 24-hour period. After 24 h, field workers returned to the participants’ homes to collect the devices and record the date and time of retrieval. Data download was performed following each collection. Minute-level data files from all visits were collated into a single longitudinal dataset for analysis. Prior to analysis, data quality control (QC) steps were performed, and readings were excluded if PM_2.5_, CO, temperature and relative humidity minute-level readings corresponded to device error flags. Following this QC, the total valid wear time was calculated, and if it was less than 50 % of the targeted wear time (24 h), that child visit was excluded from the analysis.

### Nasopharyngeal/Oropharyngeal specimen collection

Trained nurses collected combined NP/OP samples using FLOQSwabs® (Copan Diagnostics) immediately after completing the 24-hour air pollution sampling during each visit. A separate swab was used to collect specimens from the nasopharynx and oropharynx. After swabbing, the tip of the swab was placed in Skim milk–Tryptone–Glucose–Glycerol (STGG) medium. The STGG medium containing the swabs was placed in cold storage at - 20°C at the field site. The samples are then transported weekly in cold storage to a biobank and stored at -70°C until microbiology processing for isolating *S. pneumoniae*.

### *S. pneumoniae* microbiology culture

NP/OP samples were thawed on ice and up to 10 µL was inoculated on Columbia agar supplemented with 5% (v/v) defibrinated sheep blood and 5 g/L of gentamicin using the quadrant streaking method. The culture plates were then incubated overnight at 37.0°C and 5% CO2. The presumptive identification of *S. pneumoniae* was based on alpha haemolysis and colony morphology. Up to four distinct alpha-haemolytic colonies were selected and streaked on blood agar for confluent growth, and 5 µg optochin discs were placed on the inoculated plates to test for optochin susceptibility. Isolates with colonies exhibiting a zone of inhibition ≥ 14 mm were considered *S. pneumoniae*.

### Statistical Analysis

All statistical analyses were conducted using RStudio version 2024.12.0.467. Statistical tests were considered significant when p <0.05. Minute-level PM_2.5_ values at the lower detection limit (LDL) of 10 μg/m³ were substituted with LDL/√2 (= 7.07 μg/m³) as described for left-censored environmental exposure data (19). Minute-level measurements of PM_2.5,_ CO, temperature, and relative humidity were averaged over the 24-hour monitoring period to obtain child-visit level daily exposure estimates. These daily exposure estimates were then summarised across observations. Temperature and relative humidity were summarised using the arithmetic mean, while PM_2.5_ and CO were summarised using the geometric mean due to their right-skewed distributions. Arithmetic means for PM_2.5_ and CO were additionally reported to allow comparison with the WHO 2021 Air Quality Guidelines (20).

A descriptive characterisation of variations of PM_2.5_, and CO exposure by participant and household characteristics was carried out. The characteristics included: distance to the nearest main road, indoor burning of herbs or incense, floor type, household income tertile, electricity access, sex, age at visit, season, presence of a smoker in the compound and waste disposal method.

Mixed-effects logistic regression models were fitted to examine the association between exposure to PM_2.5_, CO, temperature and relative humidity and *S. pneumoniae* colonisation using the *lme4* package. *S. pneumoniae* colonisation status at each visit was set as the binary outcome. A random intercept for participants was included to account for repeated observations within participants over time. Prespecified confounders in the model included age at visit, season, and household income tertile. The confounders were based on their identification in the literature as potentially associated with *S. pneumoniae* colonisation and/or environmental exposures. PM_2.5_ and carbon monoxide (CO) concentrations were log-transformed prior to model fitting, while temperature and relative humidity were analysed on their original scales. A constant of 0.01 ppm was added to all CO values prior to log transformation to represent visits at which no measurable CO exposure was recorded. All continuous exposure variables were standardised to a mean of zero and standard deviation of one prior to model fitting to improve numerical stability and comparability across exposures. Regression coefficients were rescaled to represent ORs per IQR increase in exposure The IQR increments were: PM_2.5_, 0.66 log units; CO, 3.22 log units; temperature, 1.63°C; relative humidity, 26.8 percentage pointsThe first model (conventional model) was fitted without decomposing exposure variation yielding estimates that combine between-child and within-child sources of variation. Three sequential stages were implemented. Crude models were fitted separately for each exposure with no covariate adjustment. Single-exposure adjusted models were then fitted, each including one exposure alongside the three prespecified confounders. Finally, a mutually adjusted model was fitted including all four exposures simultaneously with the three confounders.

In the second model (Within-between decomposition model), each exposure was decomposed into two components prior to analysis; between-child and within-child. The between-child component was defined as each child’s mean exposure across all study visits, capturing their participant-level average exposure. The within-child component was defined as the deviation of each visit’s exposure value from that child’s own mean, capturing acute visit-level fluctuation above or below the child’s habitual exposure level. Both components were included simultaneously as separate predictors in each model, allowing their effects on colonisation to be estimated independently. This decomposition was applied across the same three modelling stages as the conventional analysis; crude, single exposure adjusted and mutually adjusted.

Prior to model fitting, Pearson correlations were calculated to characterise the degree of collinearity among exposures and covariates. Furthermore, variance inflation factors (VIF) were computed for all terms in the mutually adjusted models using the *car* package.

## Results

### Data analysed and participant characteristics

PM_2.5_, CO, temperature, and relative humidity exposure data were collected from 726 child visits across eight study visits. Following data quality control steps, 567 (78.1%) of the 726 child visits were included in the final dataset. The number of children sampled per visit and the number of child visits retained are presented in Table S1 (Supplementary). Of the 134 children enrolled, 132 contributed data to the final study population. The median number of monitoring visits per child in the final analysis was 5 (IQR 3–5; range 1–8). Up to 27 children (20.5%) contributed data from six or more visits.

The baseline characteristics of the 132 children in the study are presented in Table 1. There was an equal proportion of males and females (66 males and 66 females). A total of 73 households (55.3%) used firewood for cooking, whereas charcoal was used in 59 (44.7%) households. The overall median monthly household income was $62 (IQR $48–$81).

**Table 1.** Baseline characteristics of participants and their households.

| Characteristic | N = 132 |
| --- | --- |
| **Sex** |  |
| Male | 66 (50.0) |
| Female | 66 (50.0) |
| **Age at baseline, years** |  |
| Median (IQR) | 7.5 (6–9) |
| **Cooking fuel** |  |
| Wood | 73 (55.3) |
| Charcoal | 59 (44.7) |
| **Floor type** |  |
| Earthen | 15 (11.4) |
| Cement/unfinished hard floor | 88 (66.7) |
| Finished floor (tile/carpet) | 29 (22.0) |
| **Distance to main road** |  |
| <50 m | 33 (25.0) |
| 50–200 m | 52 (39.4) |
| >200 m | 47 (35.6) |
| **Household income tertiles** |  |
| Low | 44 (33.3) |
| Medium | 44 (33.3) |
| High | 43 (32.6) |
| Missing | 1 (0.8) |
| **Electricity in home** |  |
| Yes | 101 (76.5) |
| No | 30 (22.7) |
| Missing | 1 (0.8) |

### Personal PM_2.5_, CO, temperature and relative humidity distribution

The distributions of 24-hour mean personal PM_2.5_ and CO exposures were both markedly right-skewed, Figure S1, (Supplementary). PM_2.5_ exposures were generally low, with 39.3% of child-visit daily mean concentrations ≤10 µg/m³. However, a small number of elevated PM_2.5_ exposures were observed, with the 95th percentile reaching 32.3 µg/m³ and a maximum concentration of 94.2 µg/m³. CO exposures showed a more extreme right-skewed distribution, with 44.8% of child-visit daily mean concentrations <0.1 ppm and 10.6% of child visits had a 24-hour mean CO concentration of 0 ppm. Despite generally low daily mean concentrations, a small number of markedly elevated CO exposures were observed, with the 95th percentile reaching 5.3 ppm and a maximum concentration of 26.8 ppm. The PM_2.5_ and CO exposures per participant and visit is shown in Figure S2 (Supplementary)

The overall geometric mean PM_2.5_ concentration in our analysis was 12.6 µg/m³ (95% CI 12.2–13.1), and the overall geometric mean CO concentration was 0.16 ppm (95% CI 0.14– 0.19). Figure 2 illustrates the monthly variation in exposure levels. PM_2.5_ concentrations were highest in December (21.6 µg/m³) and February (19.2 µg/m³) and lowest during the mid-to-late wet season in August and September (10.2–10.7 µg/m³). CO concentrations showed a pronounced peak in March (2.32 ppm) before falling sharply in April and May (0.14-0.22 ppm), with a secondary rise during the early dry season (January-February, 1.32-1.49 ppm). Relative humidity was lowest in March (31.7%) and rose steeply from June through September (60.9-74.6%), while temperature remained relatively stable throughout the year with monthly range of 30.1-33.0 °C.

**Figure 2:**
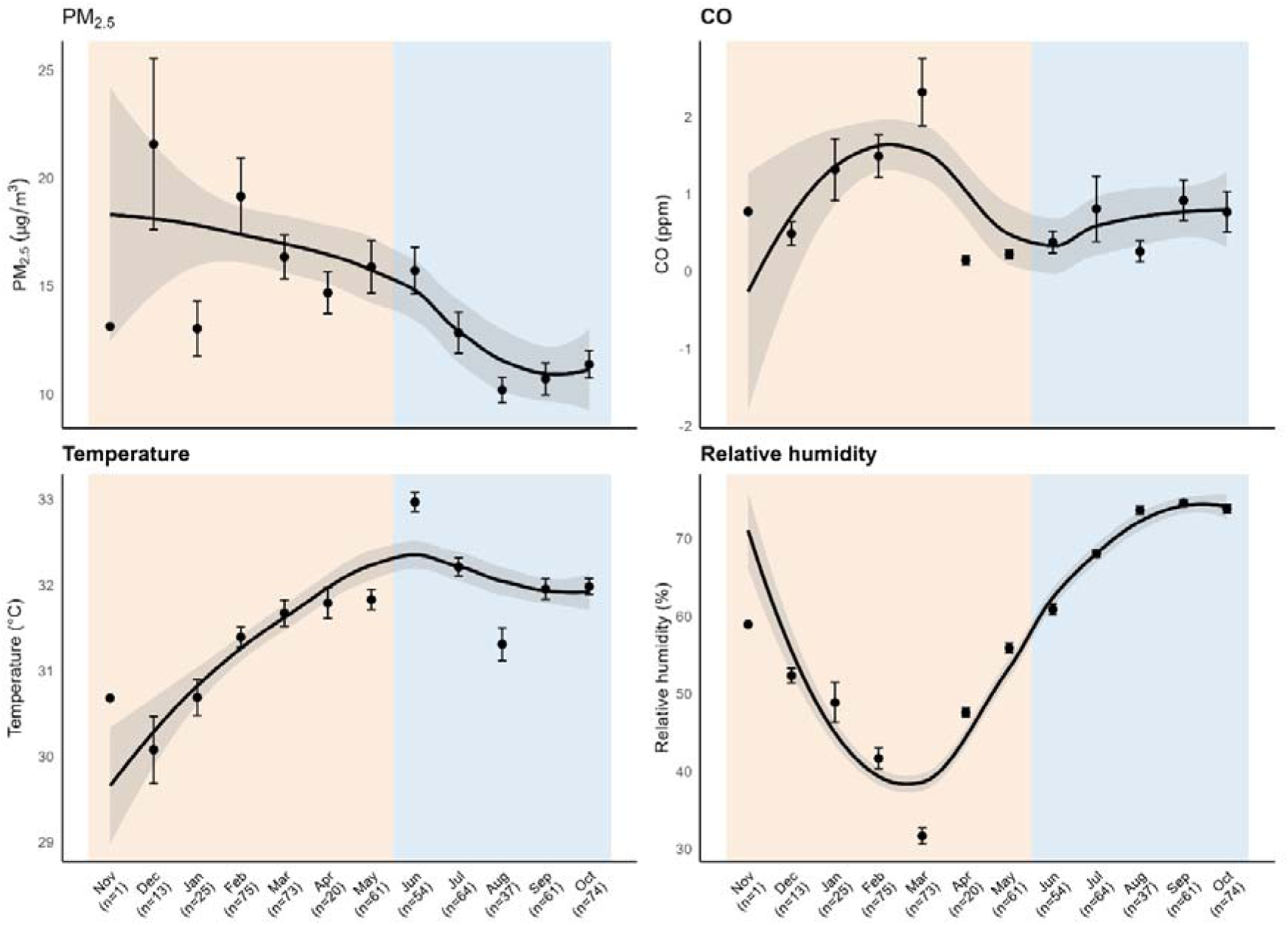
Monthly mean personal exposure to PM_2.5_, carbon monoxide, ambient temperature and relative humidity across the study period and ordered by season. Orange shading indicates dry season (November-May) and blue shading indicates the wet season (June-October).

The mean exposure by hour of the day is shown in Figure 3. Personal PM_2.5_ and CO exposures showed similarly low levels during sleeping hours (23:00–06:00), reaching their lowest levels at 05:00 for CO (0.55 ppm) and at 04:00 for PM_2.5_ (8.12 µg/m³). Distinct patterns were observed during other periods of the day. PM_2.5_ exposure increased steadily during the day, peaking at 13:00 (21.1 µg/m³). Three prominent peaks were observed for CO at 07:00 (1.05 ppm), 13:00–14:00 (1.43–1.54 ppm), and 20:00–22:00 (1.53–1.62 ppm). Temperature followed a smooth unimodal daily cycle, peaking at approximately 14:00–15:00 (35.6°C), while relative humidity showed an inverse pattern, reaching its lowest level at 15:00 (51.0%).

**Figure 3:**
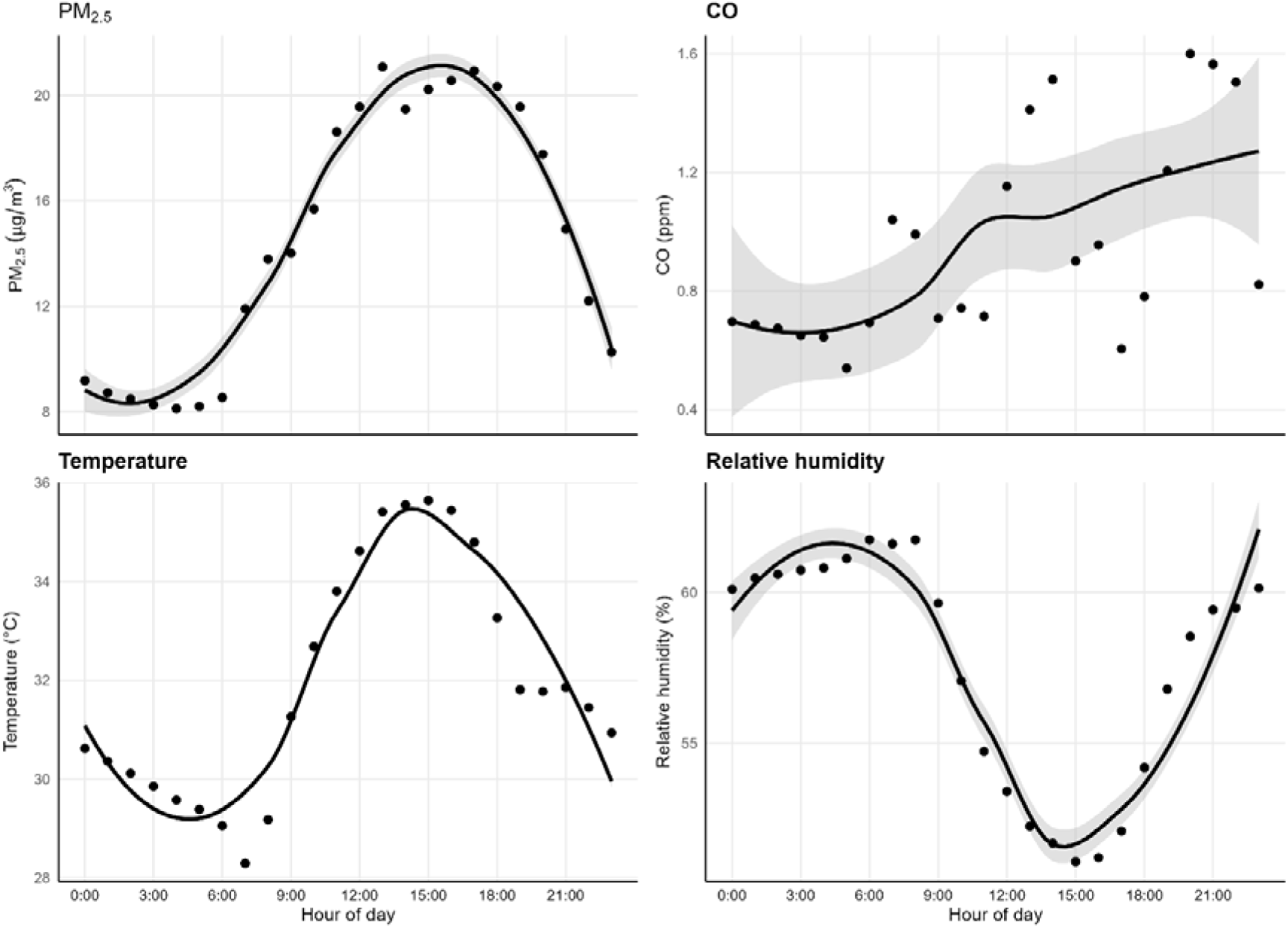
Diurnal variation in mean personal exposure to PM_2.5_, carbon monoxide, ambient temperature and relative humidity

We also compared PM_2.5_ and CO exposures with the WHO 24-hour exposure guidelines. The overall arithmetic mean PM_2.5_ exposure was 14.4 µg/m³ (SD 9.3). In 30.7% of child visits, personal PM_2.5_ exposure exceeded the WHO 2021 24-hour guideline of 15 µg/m³, Figures S3 and S4 (Supplementary). This proportion was more than twice as high in the dry season compared with the wet season (44.2% vs 18.2%). The arithmetic mean CO exposure was 0.9 ppm (SD 2.4). The WHO 24-hour CO guideline of 3.5 ppm was exceeded in 7.8% of child visits overall, with a higher exceedance rate in the dry season than in the wet season (11.6% vs 4.5%).

### Descriptive characterisation of personal PM_2.5_ and CO exposure

Children from low-income households experienced the highest PM_2.5_ (13.87 µg/m³) and CO (0.23 ppm) exposures, compared with children from medium-income households (PM_2.5_: 12.49µg/m³, CO: 0.14 ppm) and high-income households (PM_2.5_ 11.45 µg/m³ CO: 0.13 ppm), (Table 2). Children living in households with earthen floors had higher PM_2.5_ exposure (14.00 µg/m³) compared with those living in compounds with basic hard floors (12.85 µg/m³) or tiled/finished floors (11.31 µg/m³). The geometric mean PM_2.5_ exposure was higher during the dry season, 14.60 µg/m³ compared with the wet season, 11.13 µg/m³. Similarly, CO exposure was higher during the dry season, 0.23 ppm, compared with the wet season, 0.12 ppm. There was a downward trend in PM_2.5_ exposure with increasing age. A downward trend in CO exposure with increasing distance from the road was also observed. In contrast, no clear pattern was observed between PM_2.5_ exposure and distance to the road.

**Table 2.** Household and environmental characteristics and personal 24-hour geometric mean PM_2.5_ and CO exposure. GSD: Geometric standard deviation

| Characteristic | PM<sub>2.5</sub> (GSD) | CO (GSD) |
| --- | --- | --- |
| **Sex** |  |  |
| Male | 13.73 (1.66) | 0.13 (7.34) |
| Female | 11.86 (1.52) | 0.19 (7.08) |
| **Age (years)** |  |  |
| 5-8 | 14.14 (1.69) | 0.21 (6.82) |
| 9-11 | 12.23 (1.56) | 0.14 (7.79) |
| 12-14 | 11.56 (1.45) | 0.14 (6.54) |
| **Season** |  |  |
| Dry | 14.60 (1.65) | 0.23 (7.99) |
| Wet | 11.13 (1.48) | 0.12 (6.23) |
| **Cooking fuel** |  |  |
| Charcoal | 13.24 (1.74) | 0.27 (6.10) |
| Wood | 12.48 (1.54) | 0.14 (7.44) |
| **Distance to road** |  |  |
| <50 m | 12.28 (1.56) | 0.20 (6.83) |
| 50–200 m | 12.19 (1.58) | 0.16 (7.41) |
| >200 m | 13.36 (1.63) | 0.14 (7.36) |
| **Incense burning** |  |  |
| No | 14.20 (1.74) | 0.26 (7.21) |
| Yes | 12.31 (1.55) | 0.14 (7.14) |
| **Herbs burning** |  |  |
| No | 11.91 (1.57) | 0.14 (7.78) |
| Yes | 13.35 (1.61) | 0.18 (6.76) |
| **Floor type** |  |  |
| Earthen | 14.00 (1.71) | 0.17 (7.72) |
| Basic hard | 12.85 (1.59) | 0.17 (7.48) |
| Tiled/finished | 11.31 (1.50) | 0.13 (6.16) |
| **Household income** |  |  |
| Low | 13.87 (1.67) | 0.23 (6.93) |
| Medium | 12.49 (1.57) | 0.14 (6.94) |
| High | 11.45 (1.50) | 0.13 (7.67) |
| **Electricity** |  |  |
| No | 13.30 (1.69) | 0.16 (6.87) |
| Yes | 12.55 (1.58) | 0.16 (7.31) |
| **Waste disposal** |  |  |
| Household burning | 13.23 (1.66) | 0.18 (9.55) |
| Community burning | 12.22 (1.41) | 0.17 (9.08) |
| **Smoker in compound** |  |  |
| No | 12.25 (1.57) | 0.18 (7.51) |
| Yes | 13.86 (1.55) | 0.17 (12.18) |

GSD: Geometric standard deviation

### *S. pneumoniae* colonisation and personal environmental exposure

*S. pneumoniae* culture results were obtained for 97.7% (554/567) of the total samples. included in the final exposure analysis. The overall *S. pneumoniae* colonisation prevalence was 39.7% (220/554). Figure 4 shows the observed prevalence of *S. pneumoniae* colonisation across quartiles of daily environmental exposures. Colonisation prevalence increased progressively across quartiles of CO exposure, rising from 36.7% in the lowest quartile (Q1) to 43.5% in the highest quartile (Q4). In contrast, PM_2.5_ exposure showed no clear monotonic pattern, with the highest prevalence observed in the second quartile (Q2; 43.2%). In contrast, lower temperature and relative humidity quartiles generally showed higher colonisation prevalence, with the highest prevalence observed in the lowest quartile (Q1) of temperature (45.3%) as well as relative humidity (49.6%).

**Figure 4:**
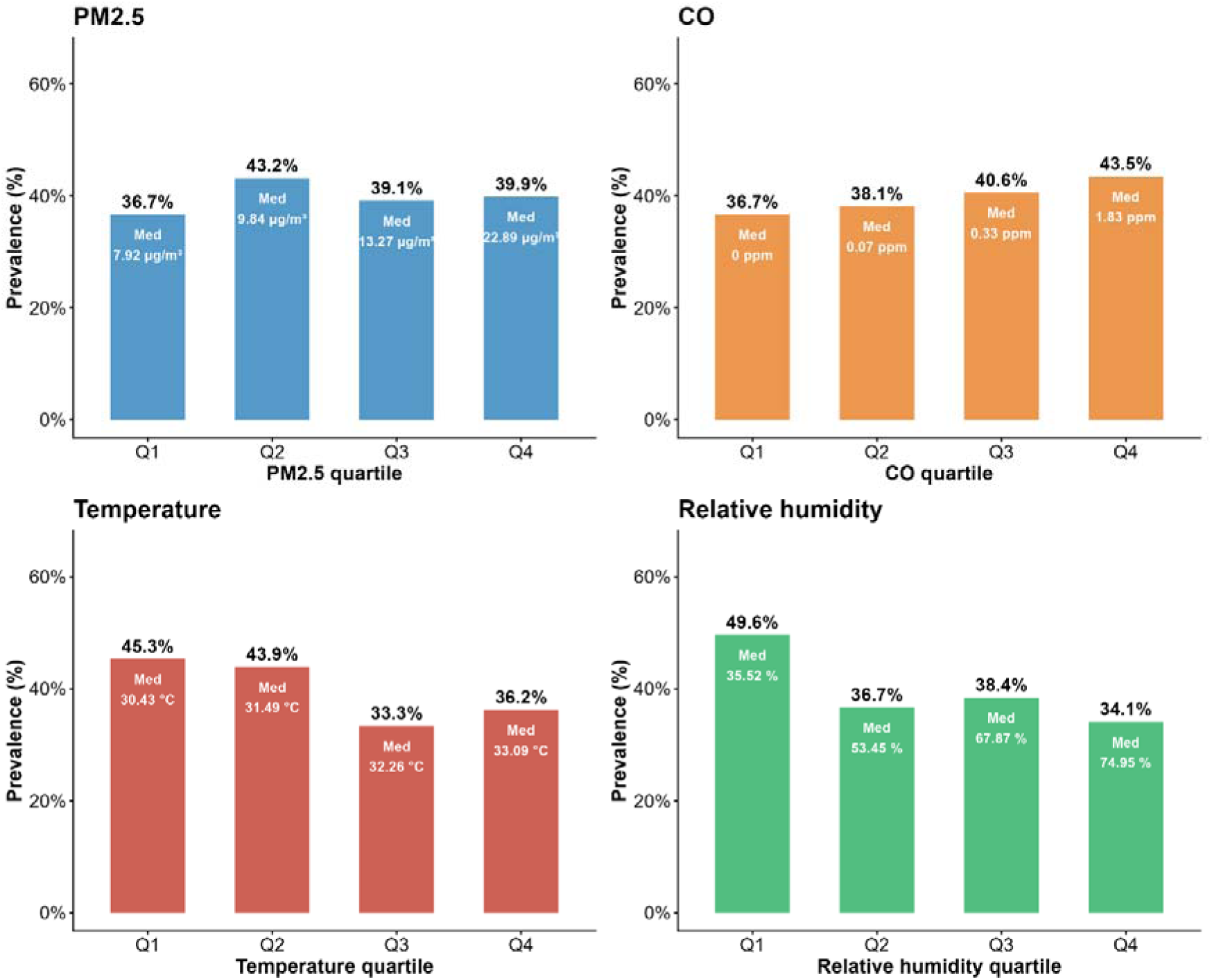
Prevalence of *S. pneumoniae* colonisation across quartiles of daily exposures of PM_2.5_, CO, Temperature, and Relative humidity. labels on top of bars indicate the *S. pneumoniae* prevalence and the labels within the bars represent the median (Med) exposure value within each quartile.

Pearson correlations among the four exposures and covariates are presented in Figure S5 (Supplementary). The highest correlation was seen between relative humidity and season. Relative humidity and season were strongly correlated (r = 0.80), consistent with the pronounced contrast between the humid wet season and the dry season. PM_2.5_ showed moderate negative correlations with relative humidity (r = −0.34). CO showed weak negative correlations with relative humidity (r = −0.22). VIF values in the mutually adjusted models were below the conventional value of 5 for all predictors (maximum VIF 3.29 for season in the within-between model), indicating that collinearity did not reach a level that would seriously compromise coefficient estimation in either model. (Table S2, Supplementary).

#### Conventional model

In the conventional Mixed-effects logistic regression models, relative humidity was significantly inversely associated with colonisation in crude analyses (OR 0.72, 95% CI 0.54–0.96; p = 0.027) (Table S3, Supplementary). Temperature showed a borderline inverse association in crude analysis (OR 0.79, 95% CI 0.61–1.02; p = 0.075). Neither PM_2.5_ (OR 1.04, 95% CI 0.81–1.34; p = 0.743) nor CO (OR 1.21, 95% CI 0.92–1.62; p = 0.204) exposure was significantly associated with *S. pneumoniae* colonisation in crude analyses of the conventional model. The significant inverse relationship between relative humidity and *S. pneumoniae* colonisation in the crude analysis was lost after adjustment for age, season, and household income in the single exposure adjusted model (OR 0.89, 95% CI 0.54–1.45; p = 0.629) and in the mutually adjusted model (OR 0.83, 95% CI 0.50–1.39; p = 0.482). Temperature also remained inversely associated with colonisation, in both the single exposure adjusted model (OR 0.84, 95% CI 0.64–1.10; p = 0.196) and mutually adjusted models (OR 0.83, 95% CI 0.62–1.09; p = 0.181) although this did not reach statistical significance. PM_2.5_ and CO were not significantly associated with colonisation in the conventional single adjusted and mutually adjusted model stages. Children from medium-income households had significantly lower odds of colonisation compared with children from low-income households across all single-exposure adjusted models (Table S4, Supplementary) and in the mutually adjusted model (OR 0.60, 95% CI 0.37–0.96; p = 0.033) (Table S5, Supplementary). A similar but non-significant reduction was observed in high-income households compared with low-income households across all four single-exposure adjusted models and in the mutually adjusted model.

#### Within-Between decomposition models

Decomposition analyses demonstrated differing patterns for between-child and within-child exposure variation (Table 3 and Figure 5). The between-child component of temperature was significantly associated with lower odds of colonisation across crude (OR 0.76, 95% CI 0.58– 0.99; p = 0.042), single adjusted (OR 0.75, 95% CI 0.57–0.98; p = 0.036), and mutually adjusted models (OR 0.74, 95% CI 0.56–0.98; p = 0.034). In contrast, no evidence of association was observed for within-child temperature fluctuations in the three model stages. Relative humidity showed a contrasting pattern. No evidence of association was observed for between-child exposure in relative humidity in the three model stages. Within-child increases in relative humidity were associated with lower odds of colonisation in the crude analysis (OR 0.70, 95% CI 0.52–0.93; p = 0.013). However, the significant association was not sustained after adjustment for season, age and income in the single adjusted and mutually adjusted models. This is consistent with the strong collinearity between relative humidity variation and season (r = 0.80)

**Table 3.**
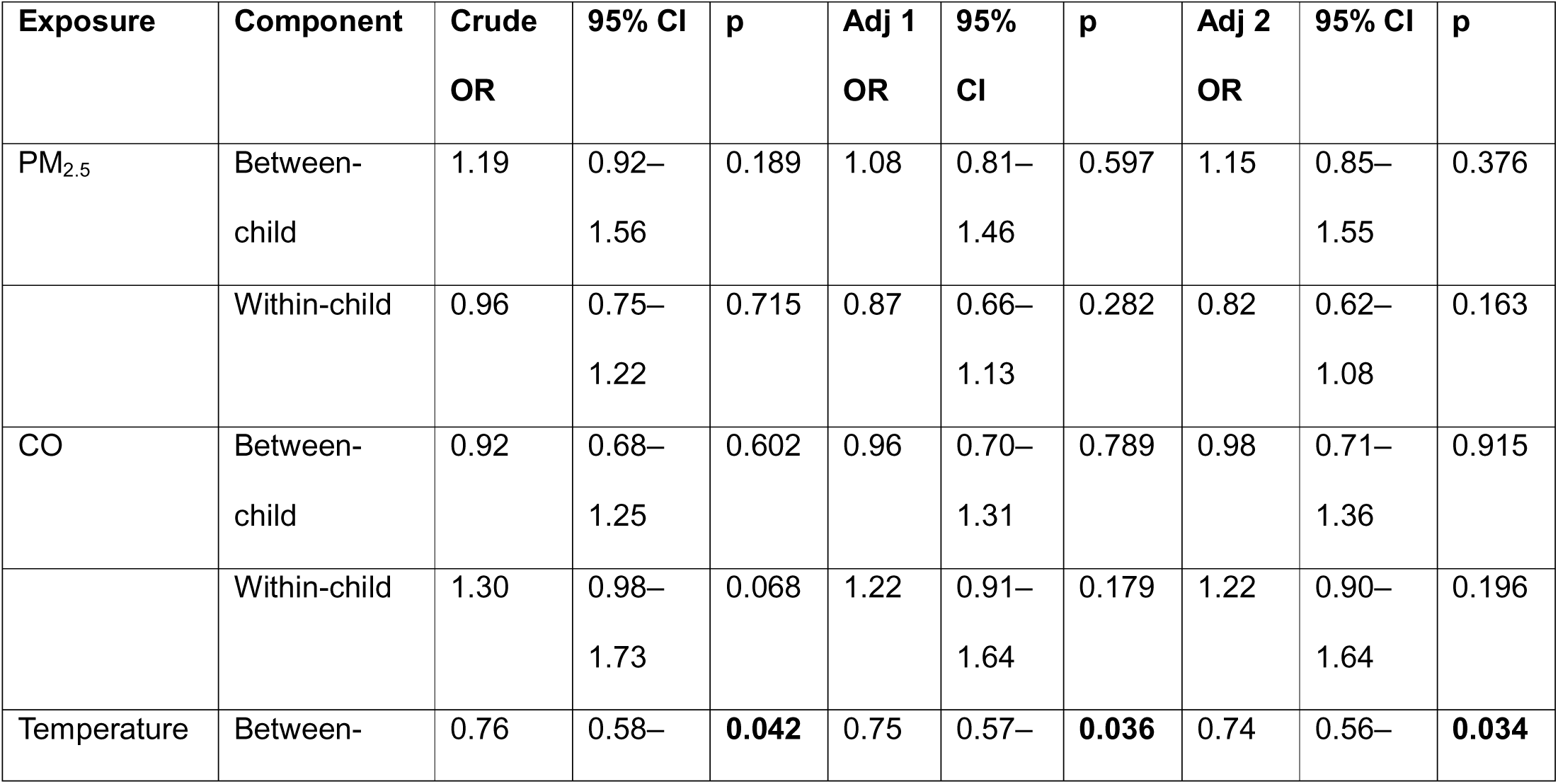
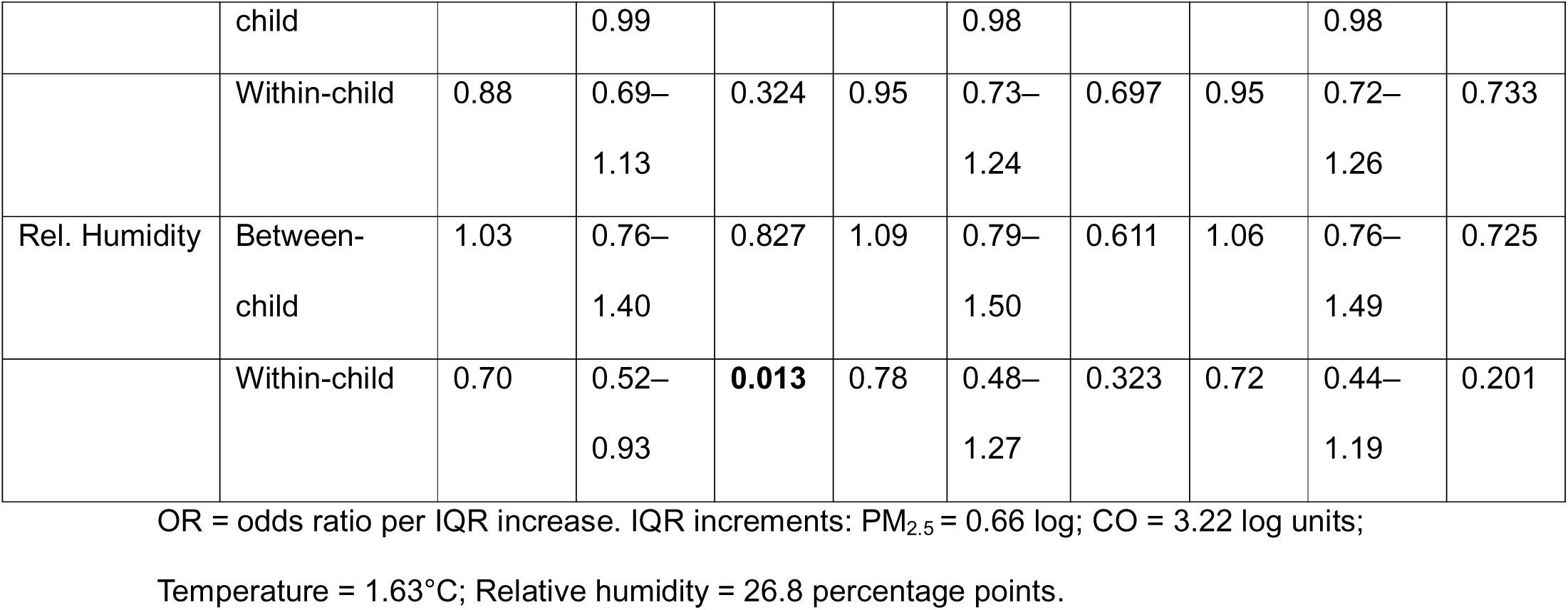
Association between environmental exposures and *S. pneumoniae* colonisation in the within-child and between-child decomposition model.

**Figure 5.**
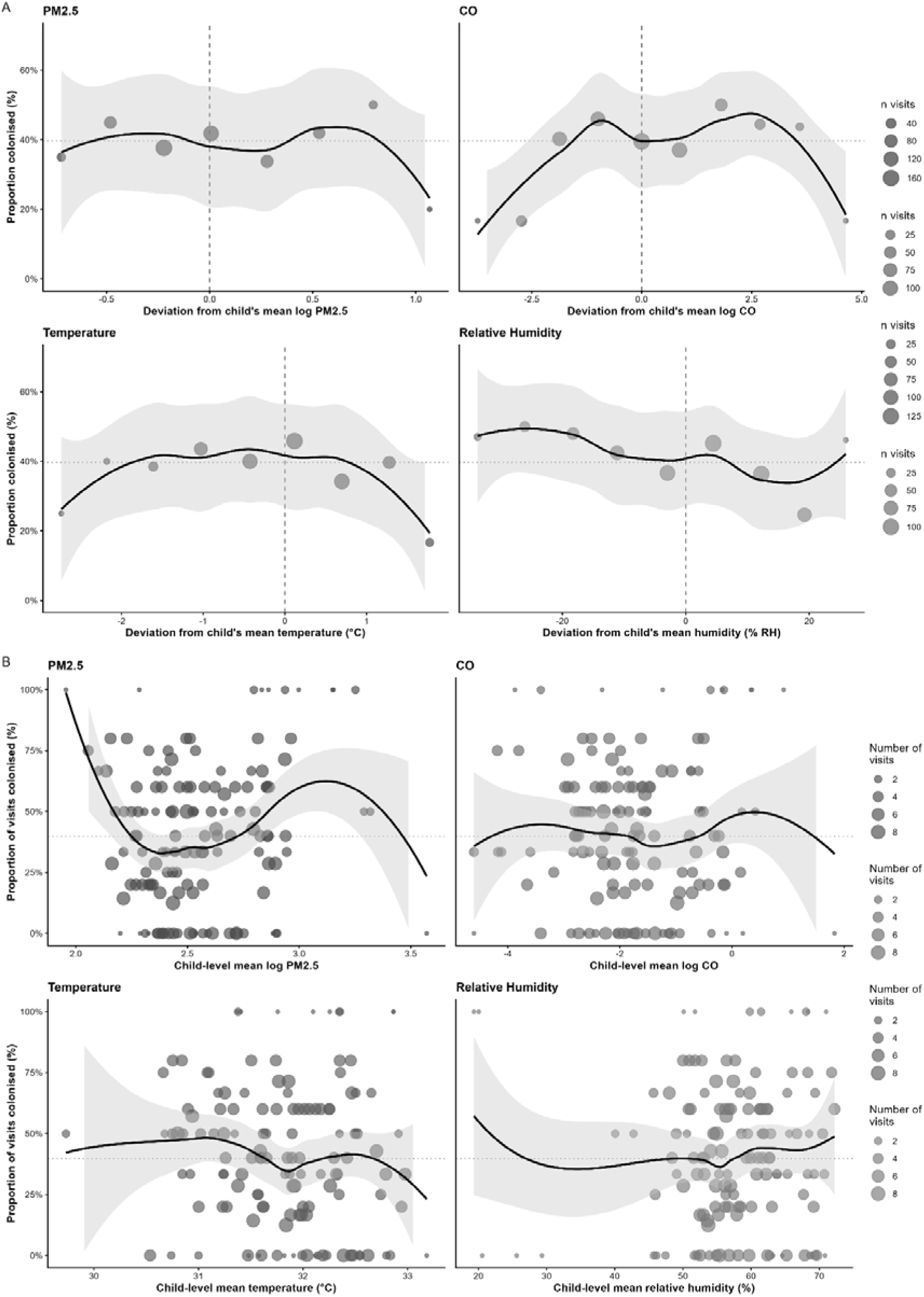
Within-child (A) and between-child (B) associations between personal exposure to PM_2.5_, CO, temperature, relative humidity and *S. pneumoniae* colonisation. Upper panel (A) show binned visit-level deviations from each child’s own exposure mean plotted against proportion colonised, where each point represents a bin of visits pooled across all children sized by number of visits. The dashed vertical line in upper panel indicates zero deviation from each child’s own mean. The dotted horizontal line indicates overall colonisation prevalence in the dataset (39.7%). Lower panel (B) show participant-level mean exposure plotted against the proportion of visits at which colonisation was detected, where each point represents one child sized by number of visits. The dotted horizontal line indicates overall colonisation prevalence in the dataset (39.7%).

No significant association was observed for PM_2.5_ and CO exposures in either between-child or within-child analyses in all three stages. CO exposure showed borderline positive within-child association in crude analyses (OR 1.30, 95% CI 0.98–1.73; p = 0.068). This possible within-child CO signal was less apparent in the conventional model, where between-child and within-child variation were combined into a single estimate.

## Discussion

In this longitudinal study of older children in rural Gambia, we combined repeated personal measurements of PM_2.5_, CO, temperature and relative humidity with microbiological assessment of *S. pneumoniae* colonisation. To our knowledge, this is the first study to report the association between personal exposure to PM_2.5_, and CO, and URT colonisation of *S pneumoniae* in children 5-14 years old, a population that has been largely overlooked in previous work. No evidence was found that personal exposure to PM_2.5_ or CO was associated with *S. pneumoniae* colonisation in either conventional mixed-effects models or within-between decomposition models. In contrast, the decomposition analysis identified a significant inverse association between the between-child component of temperature and colonisation, indicating that children who experienced higher average temperatures across follow-up had lower odds of *S. pneumoniae* colonisation. No corresponding association was observed for within-child temperature fluctuations. Relative humidity showed an inverse within-child association in crude analyses, suggesting lower odds of colonisation during visits when humidity was higher than a child’s usual level. However, this association attenuated after adjustment for season and other confounders. Overall, these findings suggest that long-term differences in temperature between children may be more relevant to pneumococcal colonisation than short-term fluctuations in temperature, while providing little evidence that PM_2.5_ or CO exposures at the levels observed in this population are important determinants of colonisation.

We focused on PM_2.5_ and CO because they are key pollutants arising from biomass combustion, which remains a dominant source of household air pollution in many rural sub-Saharan African settings. Additionally, particulate matter (PM_2.5_ and PM_10_) and CO are among the five air pollutants with the strongest evidence for public health concern according to the World Health Organisation together with ozone, nitrogen dioxide and sulfur dioxide (21). PM_2.5_ in particular is consistently associated with adverse respiratory outcomes, with both short- and long-term exposures linked to increased morbidity and mortality (22–24). Mechanistically, PM_2.5_ has been associated with epithelial injury of the upper respiratory tract (25), and may penetrate into the lungs and translocate systematically to other organs (26).

Higher levels of PM_2.5_ and CO exposure were observed during the dry season in the descriptive analysis. The Gambia experiences the dry season from November to May which includes the Harmattan period characterised by dry, dust-laden winds originating from the Sahara Desert while the rainy season spans June to October and is characterised by rainfall that could be up to 1200 mm (27, 28) during the period. The conditions experienced during the Harmattan period contribute to increased ambient particulate matter through dust transport and resuspension. In addition, dry surfaces facilitate resuspension of dust from unpaved compounds and roads during daily activities. In contrast, the wet season is associated with reduced dust resuspension due to increased ground moisture. Seasonal differences in human behaviour including increased use of biomass burning for warmth during cooler dry-season nights may also contribute to elevating both PM_2.5_ and CO exposure. Our descriptive analysis also highlights the role of socioeconomic gradients in shaping the variation of air pollution exposure. Variation in PM_2.5_ exposure by socioeconomic status are well documented globally, and our findings are consistent with this broader pattern (29–31). Although the specific reasons for the variation in our setting are not fully clear, differences in housing structure, ventilation, and flooring materials are likely contributors

We found no evidence that short-term repeated personal exposure to PM_2.5_ was associated with *S. pneumoniae* colonisation. This finding is consistent with a longitudinal birth cohort study of 158 Australians using ambient PM_2.5_ exposure (32). In that study, Takashima *et al.* (2024) followed the children for two years, examining the relationship between ambient air pollution exposure and acquisition of respiratory pathogens including *S. pneumoniae*. *S. pneumoniae* colonisation was confirmed by real-time PCR using anterior nasal swabs. PM_2.5_ concentrations were generally low (median at the time of birth 6.3 µg/m³). This null finding in a low-pollution setting using ambient exposure assessment mirrors our results despite methodological differences in exposure measurement and population setting.

In marked contrast to our findings, Deherani *et al.* (2022) reported a positive exposure-response relationship in the MSCAPE birth cohort study conducted in rural Malawi (15). Up to 485 infants were enrolled in this nested observational sub-study within a larger cookstove interventional trial, with association between PM_2.5_ exposure and *S. pneumoniae* colonisation determined at six months. The geometric mean PM_2.5_ exposure in their study was considerably higher than ours as PM_2.5_ reached 60.3 µg/m^3^ in *S. pneumoniae* colonised children and 47.0 µg/m^3^ in *S. pneumoniae* non-colonised children compared to our overall exposure of 12.6 µg/m^3^. The *S. pneumoniae* colonisation prevalence in MSCAPE was also higher at 85% compared to 40% in our study. They found that each one-decile increase in PM_2.5_ was associated with 10% higher colonisation odds after adjustment for age, sex, PCV13 vaccination, season, antibiotic use, and air pollution device runtime. The differences observed between the two studies likely reflects age specific exposure patterns and host susceptibility. Infants in rural settings would typically experience higher exposures than older children due to their proximity to maternal cooking activities. In a typical rural setting in sub-Saharan Africa, mothers are mainly responsible for cooking in the household and would strap infants on their backs while cooking (33). Older children in our cohort spent some parts of the measurement period away from household emission sources due to school activities, reducing exposure intensity. Additionally, immature immune systems in infants impair pollutant detoxification (34) and this may amplify the exposure effects of PM_2.5_ and its subsequent impact on colonisation compared to older children. Thus, both exposure magnitude and host susceptibility might be important in determining whether PM_2.5_ influences *S. pneumoniae* colonisation, with effects potentially undetectable at our lower exposure levels.

Overall exposures to CO were low in our study, consistent with findings from other studies in similar settings. This includes the Cooking And Pneumoniae study (CAPS), a cluster-randomised trial in Malawi evaluating a biomass-fuel improved cookstove intervention to reduce pneumonia incidence in children. Median exposure to CO in the intervention and control groups was 0.34 and 0.37 ppm respectively (35) in that study. No exposure-response relationship was found between CO and pneumonia incidence. Additionally, in the Ghana Randomized Air Pollution and Health Study (GRAPHS), a cluster-randomized cookstove intervention trial comparing LPG and improved biomass stoves against traditional 3-stone fires, personal median CO exposure of infants who lived in LPG compounds was 1.03 ppm (IQR: 0.3–1.91) and 0.53 ppm for the 3-stone arm (17). The exposure-response analyses showed no association between measured CO and *S. pneumoniae* colonisation in infants. On the contrary, Vanker *et al*. (2019) reported a positive association between postnatal CO exposure and *S. pneumoniae* colonisation in infants in a South African birth cohort (aRR 1.33 (95% CI 1.03–1.72) (16). Household CO exposure was measured using real-time stationary monitors during home visits and not personal monitors. The null associations between CO and *S. pneumoniae* colonisation in GRAPHS and our study despite differences in study populations, may indicate that typical personal CO exposure levels in these settings are insufficient to substantially influence colonisation risk. Methodological differences may also partly explain why the findings from Vanker and colleagues differed from the present study.

Children with higher mean temperature exposure were less likely to be colonised with *S. pneumoniae*. This finding is consistent with studies showing lower carriage during warmer periods, including evidence from Thailand among children 0-24 months old (36) and adults in Germany (37). Notably, the association was observed only for the between-child component of temperature and not for within-child fluctuations. This suggests that differences in average temperature exposure between children may be more relevant to pneumococcal colonisation than short-term changes in temperature experienced by the same child over time.

The mechanisms underlying this association remain uncertain. Experimental studies have shown that colder environmental conditions can reduce mucosal immune responses in the upper respiratory tract and this may increase susceptibility to respiratory pathogens (38). Lower temperatures have also been associated with increased transmission and persistence of several respiratory microorganisms. It is therefore plausible that children living in relatively cooler microenvironments experience conditions that favour pneumococcal acquisition or persistence. However, because the analysis was observational, the observed association cannot be assumed to reflect a direct biological effect of temperature on pneumococcal colonisation. Temperature may instead act as a marker of other environmental or behavioural factors that were not measured in this study. Further experimental studies are needed to elucidate the biological mechanisms through which temperature may influence pneumococcal carriage.

Relative humidity showed an inverse association with *S. pneumoniae* colonisation in crude analyses, and the within-between decomposition model further suggested that visits with higher-than-usual humidity were associated with lower odds of colonisation. However, this association was attenuated after adjustment for season, indicating that the effects of humidity are difficult to disentangle from broader seasonal influences. In The Gambia, pneumococcal colonisation has previously been shown to be higher during the dry season, (39) which is characterised by lower humidity. The findings in this study are consistent with this pattern and suggest that short-term increases in humidity may be associated with reduced colonisation risk. However, because humidity is strongly correlated with season, it remains unclear whether the observed association reflects a direct effect of humidity itself or other seasonal processes, such as changes in crowding, social mixing, respiratory viral circulation, or rainfall-related environmental conditions. These findings highlight the challenges of separating the effects of closely correlated environmental exposures.

Notably, household income was a consistent predictor of *S. pneumoniae* colonisation with higher odds observed among children from low-income households in the conventional mutually adjusted model. This suggests that characteristics associated with low-income households such as crowding, living conditions may play an important role in shaping colonisation risk.

This study has several important strengths, including leveraging a longitudinal design with repeated measurements of both personal environmental exposures and *S. pneumoniae* colonisation outcome in an age group that has not been studied. This allowed for a robust assessment of within-child variation over time in contrast to earlier studies that relied on limited timepoints or indirect exposure measures.

Our study is not without limitations. Air pollution exposures were measured over 24-hour periods and therefore represent short-term exposure rather than cumulative exposure histories that may be more relevant for colonisation processes. Although repeated measurements partially mitigate this limitation, the study cannot exclude potential effects of longer period exposure measurements. Secondly, colonisation was assessed as a binary outcome, which lacks sensitivity to detect changes in bacterial density.

## Abbreviations

PM_2.5_: Particulate matter
CO: Carbon monoxide
ARIs: Acute respiratory infections
GRAPHS: Ghana Randomized Air Pollution and Health Study
CAPS: Cooking And Pneumoniae study
PATS+: Particle and temperature sensor
LDL: lower detection limit
ppm: Parts per million
PICA: Platform for Integrated Cookstove Assessment
STGG: Skim milk–Tryptone–Glucose–Glycerol
VIF: Variance inflation factors

## Ethics approval and consent to participate

The study was approved by the Joint Medical Research Council Unit The Gambia at the London School of Hygiene and Tropical Medicine and the Government of The Gambia ethics committee (Ethics ref: 26449). Written informed consent was obtained from the parents or guardians of all participants prior to enrolment. A copy of the participant information sheet and consent form were provided to each guardian. The content was interpreted in the local language for guardians who were not fluent in English. All procedures were performed in accordance with the relevant guidelines and regulations.

## Consent for publication

Not applicable

## Availability of data and materials

The data that supports the findings of this study are available upon request, given the requirement for participant confidentiality under the informed consent obtained for this study. Researchers may apply for access through the Gambia Government/MRC Joint Ethics Committee, facilitated by MRC Unit The Gambia https://www.lshtm.ac.uk/research/units/mrc-gambia through Martin Antonio, or Dam Khan. https://www.lshtm.ac.uk/research/units/mrc-gambia

## Competing interests

The authors declare that they have no competing interests

## Funding

This work is supported by the U.S. National Institutes of Health (NIH), Office of the Director (OD), the National Institute of Environmental Health Sciences (NIEHS), and the National Human Genome Research Institute (NHGRI) grant number U54HG009824.

## Authors’ contributions

BKA, MA and MPN conceived and designed the study. PN, AW and MAK managed sample collection and data tracking. PET, SAT and DK performed laboratory processing. DK and NIM conducted the data analysis. DK prepared the figures, generated tables and wrote the initial draft of the Manuscript. MPN, NIM, and MA edited and revised the manuscript.

## Acknowledgements

We express our sincere gratitude to study participants and their parents or guardians for their time and cooperation, and to the ‘Alkali’ and local village authorities for facilitating the study.

## Notes

### Competing Interest Statement

The authors have declared no competing interest.

