## Supplementary Information for "Longitudinal personal environmental exposure and *Streptococcus pneumoniae* colonisation in a cohort of children 5-14 years old in rural Gambia"

Table S1. Personal exposure monitoring across study visits showing child-visits monitored and retained after quality control

| **Visit** | **Child-visits monitored** | **Child-visits retained after QC n (%)** |
| --- | --- | --- |
| 1 | 62* | 61 (98.4) |
| 2 | 120 | 72 (60.0) |
| 3 | 101 | 58 (57.4) |
| 4 | 86 | 34 (39.5) |
| 5 | 102 | 100 (98.0) |
| 6 | 88 | 88 (100.0) |
| 7 | 96 | 89 (92.7) |
| 8 | 71 | 65 (91.5) |
| Total | 726 | 567 (78.1) |

* Device availability was limited during Visit 1. Therefore only 62 of the consented 134 study participants were monitored during this visit.

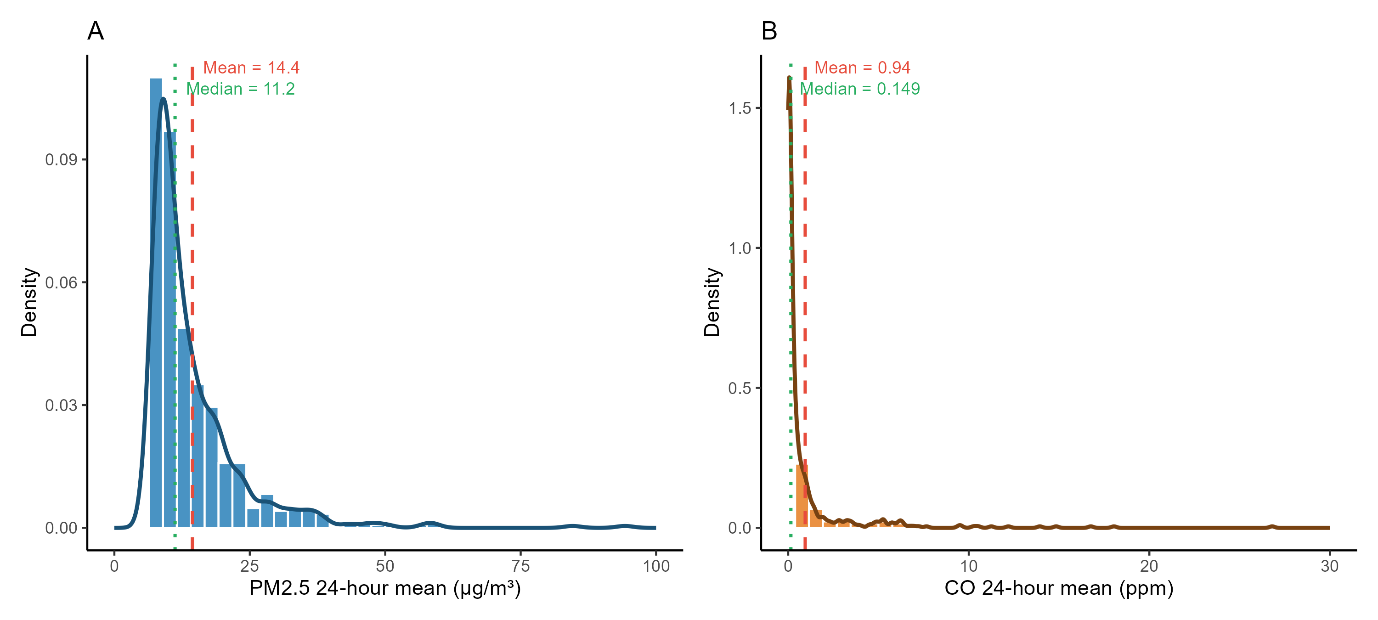

Figure S1: Distribution of 24-hour mean personal PM_2.5_ (A) and CO (B) exposures across all child-visits. The Red dashed line indicates the arithmetic mean and the green dotted line indicates the median.

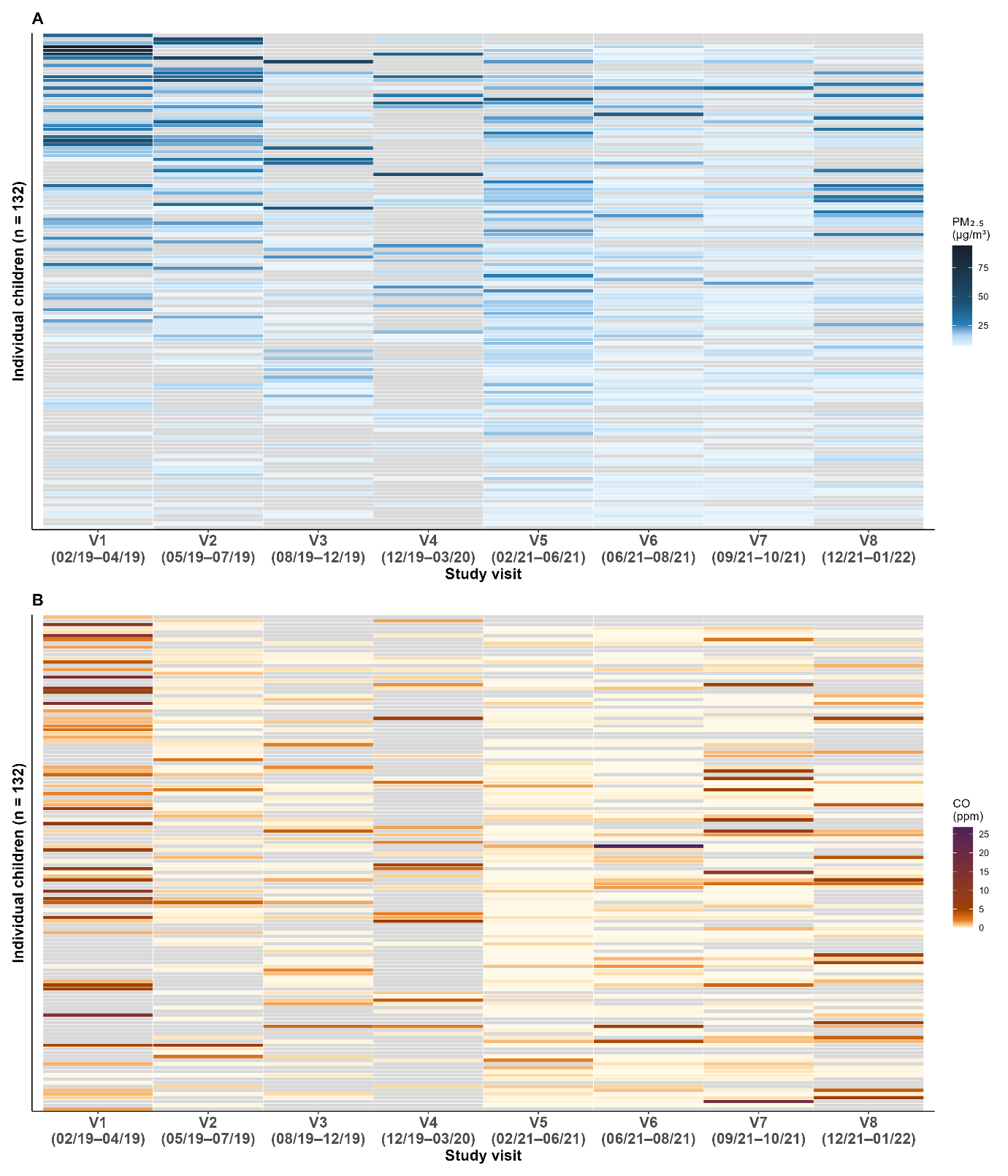

Figure S2: Longitudinal air pollution exposure profiles for individual children by study visit

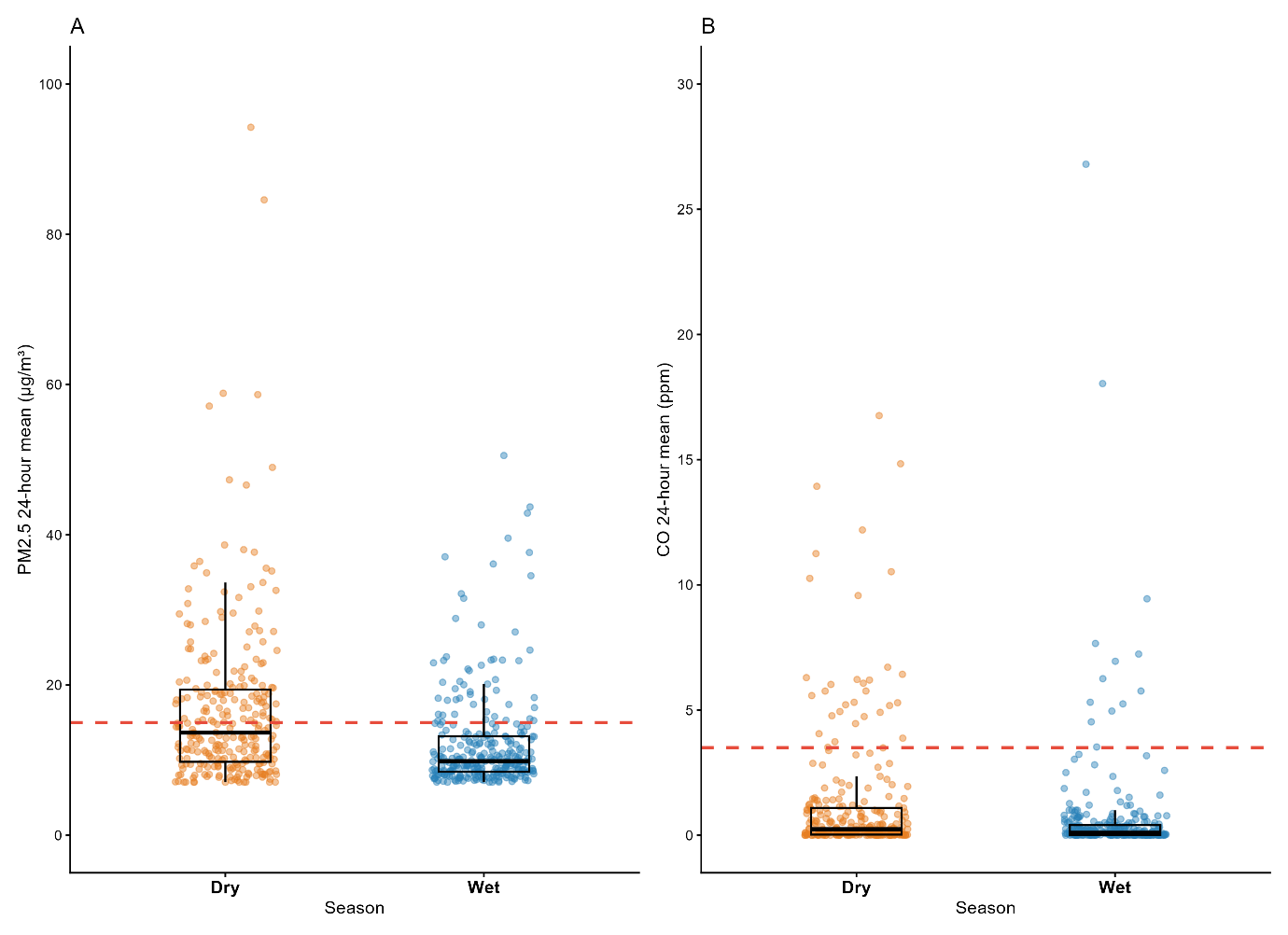

Figure S3: Individual 24-hour mean personal PM_2.5_ (A) and CO (B) exposures by season. Each point represents one child-visit. Red dashed line indicates the WHO 24-hour guideline (PM_2.5_ :15 µg/m³ and CO: 3.5 ppm).

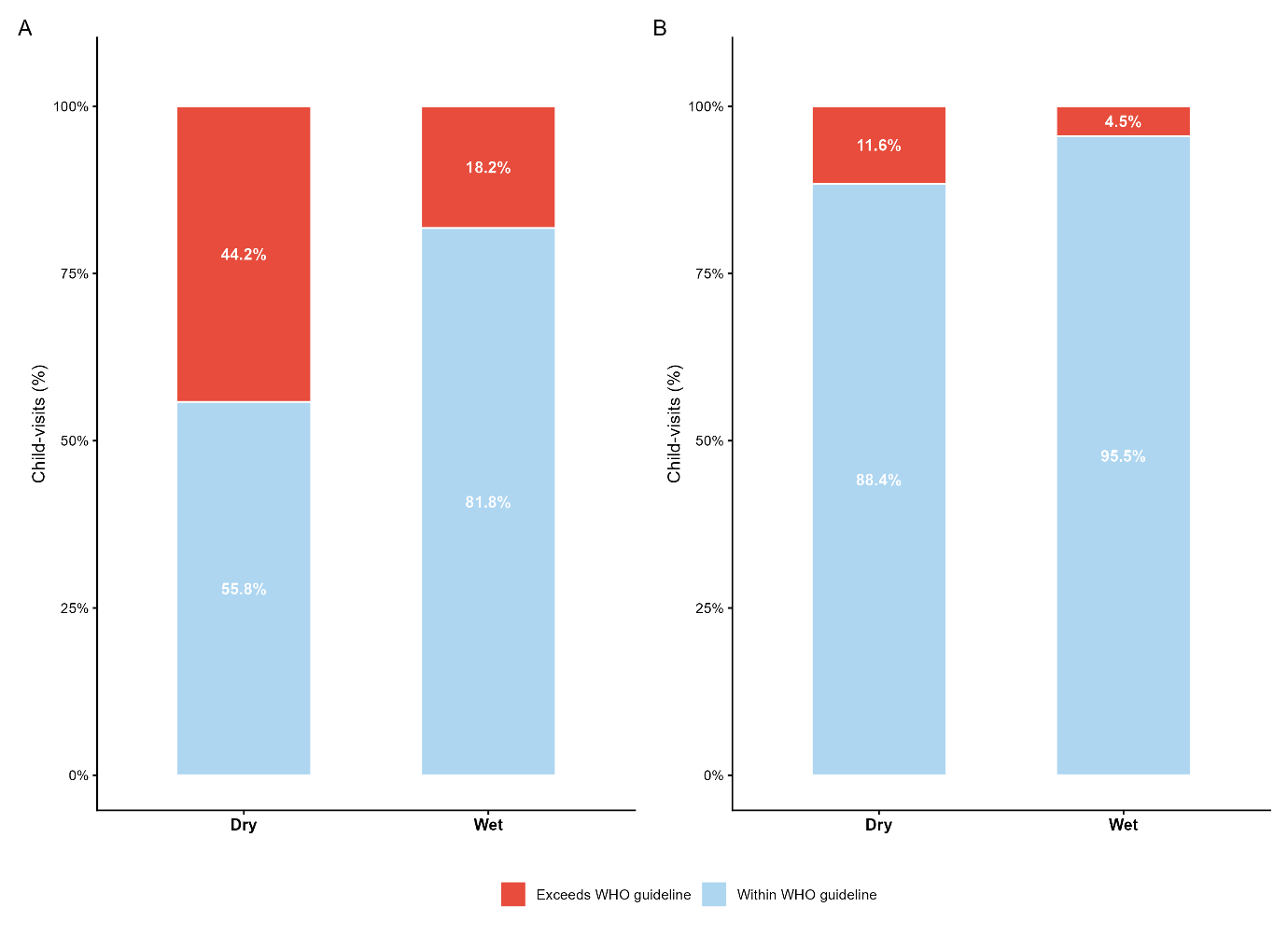

Figure S4: Proportion of child-visits exceeding WHO 24-hour air quality guidelines by season. A: PM_2.5_ exposure exceeding 15 µg/m³. B: CO exposure exceeding 3.5 ppm.

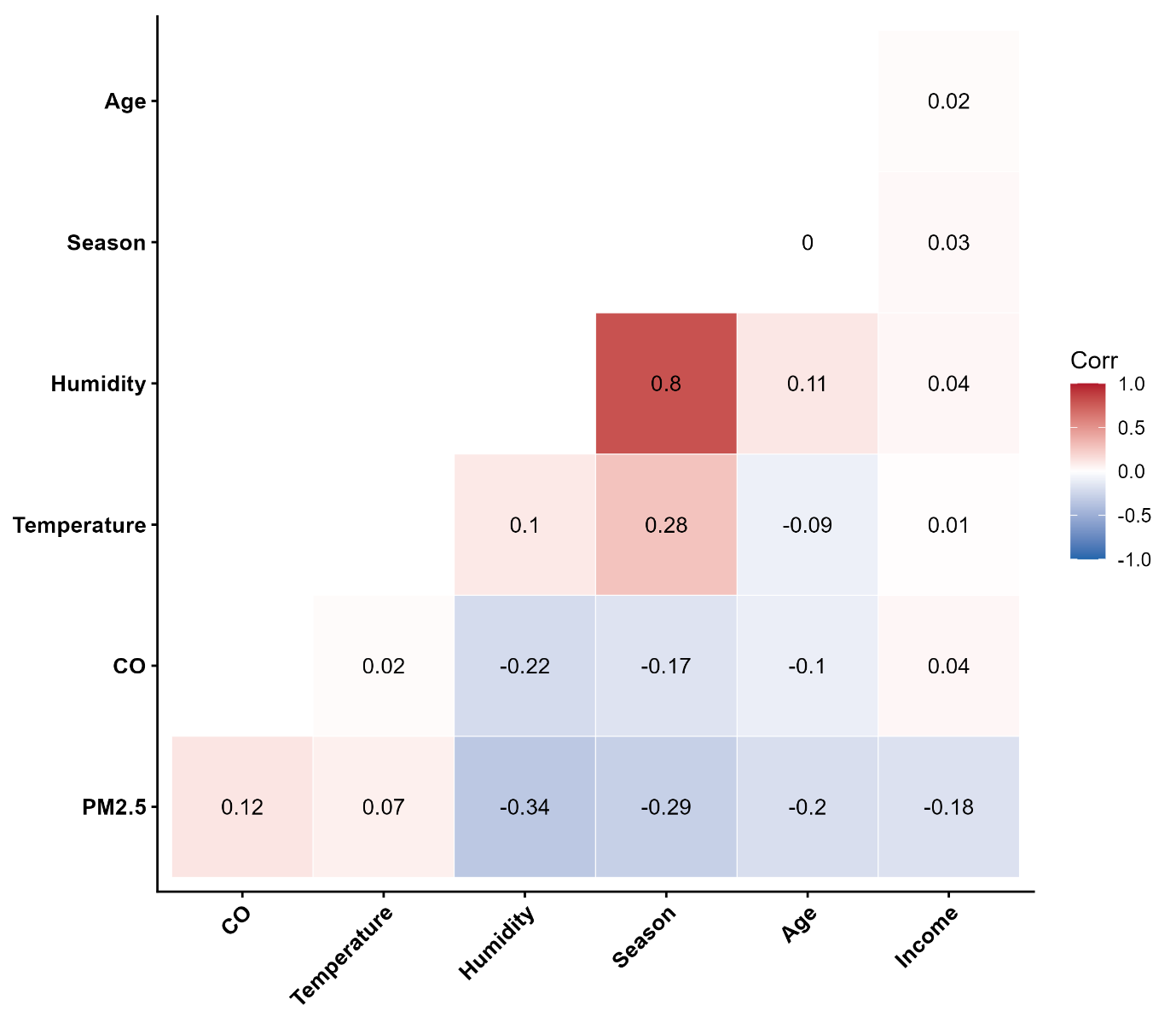

Figure S5. Correlation matrix of personal environmental exposures and covariates. Colour intensity reflects the magnitude of the r coefficients. Binary season is coded as 0 = dry season and 1 = wet season. Household income is analysed across ordinal tertiles (1 = lowest, 3 = highest).

Table S2. Variance inflation factors (VIF) for mutually adjusted Models

| **Covariates** | **VIF** |
| --- | --- |
| **Conventional mutually adjusted model** |  |
| PM_2.5_ | 1.25 |
| CO | 1.07 |
| Temperature | 1.15 |
| Relative Humidity | 3.08 |
| Age | 1.08 |
| Season | 3.18 |
| Income tertile† | 1.01 |
| **Within-Between mutually adjusted model** |  |
| PM_2.5_ - between-child | 1.25 |
| PM_2.5_ - within-child | 1.23 |
| CO - between-child | 1.11 |
| CO - within-child | 1.07 |
| Temperature - between-child | 1.05 |
| Temperature - within-child | 1.15 |
| Humidity - between-child | 1.25 |
| Humidity - within-child | 3.08 |
| Age | 1.14 |
| Season | 3.29 |
| Income tertile† | 1.05 |

† generalised VIF standardised by degrees of freedom is reported.

Table S3***.*** Association between environmental exposures and *S. pneumoniae* colonisation in the conventional mixed effects logistic regression models.

| **Exposure** | **Crude OR** | **95% CI** | **p** | **Single exposure adjusted model OR** | **95% CI** | **p** | **Mutually adjusted model OR** | **95% CI** | **p** |
| --- | --- | --- | --- | --- | --- | --- | --- | --- | --- |
| PM_2.5_ | 1.04 | 0.81–1.34 | 0.743 | 0.90 | 0.69-1.19 | 0.47 | 0.91 | 0.68-1.20 | 0.494 |
| CO | 1.21 | 0.92–1.62 | 0.204 | 1.15 | 0.85-1.56 | 0.348 | 1.16 | 0.86-1.58 | 0.332 |
| Temperature | 0.79 | 0.61–1.02 | 0.075 | 0.84 | 0.64-1.10 | 0.196 | 0.83 | 0.62-1.09 | 0.181 |
| Rel. humidity | **0.72** | **0.54–0.96** | **0.027** | 0.89 | 0.54-1.45 | 0.629 | 0.83 | 0.50-1.39 | 0.482 |

OR = odds ratio per IQR increase for exposures; IQR increments: PM_2.5_ = 0.66 log units; CO = 3.22 log units; temperature = 1.63°C; relative Humidity = 26.8 percentage points.

Table S4. Covariates in the single exposure adjusted models.

| Covariates | **PM_2.5_ model** | | **CO model** | | **Temperature model** | | **Relative humidity model** | |
| --- | --- | --- | --- | --- | --- | --- | --- | --- |
| **Conventional model** | | | | | | | | |
|  | OR (95% CI) | p | OR (95% CI) | p | OR (95% CI) | p | OR (95% CI) | p |
| Age (per year) | 0.91 (0.75-1.1) | 0.335 | 0.93 (0.77-1.13) | 0.48 | 0.91 (0.75-1.1) | 0.333 | 0.93 (0.77-1.13) | 0.484 |
| Season: Wet vs Dry | 0.67 (0.46-0.97) | **0.034** | 0.72 (0.5-1.03) | 0.072 | 0.75 (0.52-1.08) | 0.123 | 0.81 (0.44-1.46) | 0.476 |
| Income: Medium vs Low | 0.62 (0.38-0.99) | **0.044** | 0.61 (0.38-0.98) | **0.04** | 0.62 (0.39-0.99) | **0.046** | 0.62 (0.39-1) | **0.048** |
| Income: High vs Low | 0.64 (0.4-1.04) | 0.074 | 0.65 (0.41-1.06) | 0.082 | 0.67 (0.42-1.06) | 0.089 | 0.67 (0.41-1.07) | 0.094 |
| **Within-Between model** | | | | | | | | |
| Age (per year) | 0.92 (0.76-1.12) | 0.413 | 0.94 (0.78-1.14) | 0.535 | 0.9 (0.74-1.09) | 0.268 | 0.94 (0.78-1.14) | 0.542 |
| Season: Wet vs Dry | 0.65 (0.45-0.95) | **0.026** | 0.72 (0.5-1.03) | 0.075 | 0.72 (0.5-1.05) | 0.087 | 0.86 (0.47-1.58) | 0.63 |
| Income: Medium vs Low | 0.64 (0.4-1.03) | 0.064 | 0.63 (0.39-1.01) | 0.057 | 0.62 (0.39-0.99) | **0.045** | 0.62 (0.38-0.99) | **0.044** |
| Income: High vs Low | 0.7 (0.42-1.16) | 0.167 | 0.67 (0.41-1.07) | 0.094 | 0.67 (0.42-1.07) | 0.094 | 0.65 (0.4-1.05) | 0.076 |

Table S5. Covariates in the mutually adjusted models.

| **Covariate** | **OR** | **95% CI** | **P** |
| --- | --- | --- | --- |
| **Conventional model** |  |  |  |
| Age (per year) | 0.92 | 0.75–1.12 | 0.386 |
| Season: Wet vs Dry | 0.87 | 0.46–1.65 | 0.677 |
| Income: Medium vs Low | **0.60** | **0.37–0.96** | **0.033** |
| Income: High vs Low | 0.64 | 0.40–1.05 | 0.075 |
| **Within-Between decomposition model** |  |  |  |
| Age (per year) | 0.94 | 0.77–1.15 | 0.524 |
| Season: Wet vs Dry | 0.88 | 0.46–1.69 | 0.708 |
| Income: Medium vs Low | 0.64 | 0.39–1.03 | 0.066 |
| Income: High vs Low | 0.72 | 0.43–1.20 | 0.211 |
